# Association between the physical work environment and work functioning impairment while working from home under the COVID-19 pandemic in Japanese workers

**DOI:** 10.1101/2021.03.23.21254207

**Authors:** Makoto Okawara, Tomohiro Ishimaru, Seiichiro Tateishi, Ayako Hino, Mayumi Tsuji, Kazunori Ikegami, Masako Nagata, Shinya Matsuda, Yoshihisa Fujino, for the CORoNaWork project

**Affiliations:** Department of Environmental Epidemiology, Institute of Industrial Ecological Sciences, University of Occupational and Environmental Health, Japan; Department of Occupational Medicine, School of Medicine, University of Occupational and Environmental Health, Japan; Department of Mental Health, Institute of Industrial Ecological Sciences, University of Occupational and Environmental Health, Japan; Department of Environmental Health, School of Medicine, University of Occupational and Environmental Health, Japan; Department of Work Systems and Health, Institute of Industrial Ecological Sciences, University of Occupational and Environmental Health, Japan; Department of Occupational Health Practice and Management, Institute of Industrial Ecological Sciences, University of Occupational and Environmental Health, Japan; Department of Preventive Medicine and Community Health, School of Medicine, University of Occupational and Environmental Health, Japan

**Keywords:** Environment and Public Health, workplace, Posture, presenteeism, Work Performance, COVID-19

## Abstract

**Objective:** This study examined the relationship between the physical work environment and work functioning impairment while working from home in the context of rapid changes associated with the COVID-19 pandemic.

**Methods:** This cross-sectional study of internet monitors was conducted between December 22 and 26, 2020. Of a total of 33,302 participants, 5,760 who worked from home at least 1 day a month, excluding those who met the exclusion criteria, were included in the analysis. A binary subjective assessment of the physical work environment while working from home was used as an exposure factor. We examined 9 items related to the physical work environment, including level of illuminance and use of suitable desks and chairs, traditionally recommended for health and safety management when working at a desk. The number of non-conformities to 7 items was also used as an exposure factor. The presence of severe work functioning impairment was measured using the Work Functioning impairment Scale (WFun), a self-reported outcome measure of the degree of work functioning impairment. Odds ratios of severe work functioning impairment were estimated using mixed-effects logistic regression analysis with the prefecture of residence as a random effect.

**Results:** Multivariate analysis showed that the odds ratio of severe work functioning impairment was significantly higher among those who indicated “No” to all recommended items except for “I work at a desk/chair for office use.” The highest odds ratio of work functioning impairment was associated with a “No” response to “There is enough light to do my work” (aOR: 2.02, 95%CI: 1.73–2.35, p<0.01). Our results also suggest the presence of a dose-response relationship between the number of non-conformities to recommendations for work environments while working from home and work functioning impairment.

**Conclusions:** Our findings suggest that it is important for both companies and individual workers to create a work environment that prevents negative health outcomes and improves productivity while working from home.

## Introduction

The expansion of telecommuting brought about by the COVID-19 pandemic is one of the most marked changes to the way people work around the world in decades. In February 2020, the Japanese government issued a basic policy describing countermeasures against COVID-19, in which it included a recommendation for companies to implement telework to prevent the spread of infection (1). Furthermore, as the disease continued to spread throughout Japan, the government declared a state of emergency in April 2020, requesting people to refrain from going out, and further promoting telework (2). As a result, the number of companies adopting telecommuting and the percentage of workers working from home have increased rapidly (3). Further, even after the state of emergency ended, more and more companies continued to employ coronavirus countermeasures, mixing going to work and working from home, based on the situation in each region (4).

Due to the unexpected emergence of COVID-19, neither companies nor workers were able to sufficiently prepare their home environments for conducting work. While work-from-home systems had been discussed as a way to counter overwork and promote work-life balance in Japan, few effective efforts had been made (5). However, with the COVID-19 pandemic, companies were forced to rapidly adopt telework at the request of the national and local governments. As a result, many workers have had to work from home in environments that are not fully equipped with the necessary systems or facilities, and that are suboptimal for work execution. In 2019, about 20% of companies in Japan had adopted telework. In contrast, in March 2020, 24% of companies in Tokyo had adopted telework, jumping to about 57% in January 2021 (4,6).

Working from home is expected to have an impact on workers’ health and work productivity. Working in environments that are not properly designed and working with poor posture can affect the musculoskeletal system (7,8,9). In addition, there are concerns that individuals are working longer due to insufficient management of working hours. Further, reduced opportunities for direct communication can make it difficult for individuals to receive support from superiors and colleagues, and for managers to manage the situation. These factors have been linked to feelings of loneliness, isolation, and depressed mood (10,11,12).

Workers’ physical work environment while working from home is important for their health and work productivity. There are several basic recommendations for the working-from-home environment: have enough space to work; control the illumination of text, keyboard surfaces, and displays; prevent glare from displays; provide windows and other ventilation equipment; maintain a constant temperature and humidity in the room; reduce noise; and prepare necessary equipment, including desks and chairs, to enable a good work posture and provide an effective workspace (13,14,15).

However, because working from home has rapidly become the new way of working in Japan during the COVID-19 pandemic, it is not known how the physical work environment while working from home affects worker’s health and productivity. In fact, many companies have no control over workers’ work environment while they are working from home. A previous survey reported that, of the 1,256 companies in Japan that had introduced telework, 75.8% indicated that they did not check any components of workers’ physical work environment, such as air conditioning, lighting, and desk and chair condition, while they were working from home (16). Working from home in an environment that is not properly maintained can cause work functioning impairment through worker’s health problems.

The purpose of this study was to investigate the relationship between the physical work environment while working from home and work functioning impairment in the context of rapid changes associated with the COVID-19 pandemic.

## Methods

This cross-sectional study was conducted using data from the baseline survey of the Collaborative Online Research on the Novel-coronavirus and Work (CORoNaWork) Project, a prospective cohort study that used a questionnaire survey of Internet monitors to investigate the health of workers during the COVID-19 pandemic (17). This study was conducted with the approval of the Ethics Committee of the University of Occupational and Environmental Health, Japan (Approval No. R2-079).

The survey was conducted between December 22 and 26, 2020. A total of 33,302 workers between the ages of 20 and 65 years at the time of the survey were included, and sampling was designed such that participants’ sex and occupation (office workers and non-office workers) were balanced by region of residence, based on the cumulative infection rate of COVID-19. Excluding those who were deemed to have provided inappropriate responses, the total number of participants in this study was 27,036. Details of the exclusion criteria are described in the protocol (17). Briefly, we excluded those who had extremely short response times, were shorter than 140 cm tall, weighed less than 30 kg, or provided inconsistent responses to multiple identical questions. In this study, workers who were working from home at least 1 day a month were included in the analysis.

### Assessment of physical work environment while working from home

A binary subjective assessment of the physical work environment while working from home was used as an exposure factor. The following nine items were examined: “There is a place/room where I can concentrate,” “There is enough light to do my work,” “There is enough space on my desk to work,” “There is enough space at my feet,” “The temperature and humidity in the room are comfortable,” “The environment is quiet,” “I work at a desk/chair for office use,” “I work at a desk/chair not for office use,” “I work at a coffee table or kotatsu.” We included the final item “I work at a coffee table or kotatsu” because sitting on the floor at a low table such as a coffee table or kotatsu (a low table equipped with a heater) to eat or do light work is a common way of life in Japan.

Additionally, we examined the number of non-conformities to seven of the nine recommended items for work environments; we excluded “I work at a desk/chair for office use” and combined responses to the items “I work at a desk/chair not for office use” and “I work at a table/kotatsu.” Thus, non-conformity was indicated by “No” responses to the following items: “There is a place/room where I can concentrate,” “There is enough light to do my work,” “There is enough space on my desk to work,” “There is enough space at my feet,” “The temperature and humidity in the room are comfortable,” “The environment is quiet,” and “I work at a desk/chair not for office use” or “I work at a coffee table or kotatsu.” The number of non-conformities was stratified into five categories (0 items, 1-2 items, 3-4 items, 5-6 items, and 7 items) and used as an exposure factor.

### Assessment of work functioning impairment and other covariates

The presence or absence of severe work functioning impairment was measured using the Work Functioning impairment Scale (WFun) and used as the primary outcome. WFun is a self-reported outcome measure of the degree of work functioning impairment developed based on the Rasch model and validated according to Consensus‐based Standards for the selection of health Measurement Instruments (COSMIN) (18). Subjects provide responses to a total of seven items, including “I haven’t been able to behave socially” and “I have felt that my work isn’t going well,” on a five-point scale, and the total score indicates the degree of work functioning impairment. The total score ranges from 7 to 35, and a score of 21 or higher is defined as severe work functioning impairment based on the results of interviews with occupational health nurses in a previous study (19). While the original WFun contains seven items, we used a six-item version, from which the scores can be equivalently converted to those of the original version based on the Rasch model.

For socioeconomic factors, we examined the following items: sex, age, education (junior high school; high school; vocational school, junior college, or technical college; university or graduate school), marital status (married; divorced/bereaved; never married), job type (mainly desk work; Jobs mainly involving interpersonal communication; mainly physical work), equivalent income (household income divided by the square root of household size), company size (total number of employees in the company where the respondent mainly works [self-employed answered one]).

For work-related factors, we examined the following items: frequency of working from home (at least 4 days a week; at least 2 days a week; at least 1 day a week; at least 1 day a month; hardly ever), change in working hours compared to pre-2019 (increased; no change; decreased), one-way commute time.

As the community-level factor, we used the cumulative incidence of COVID-19 infection in the prefecture of residence one month before the survey. Data were collected from the websites of public institutions.

### Statistical analysis

Age and WFun scores are presented as continuous variables using median and interquartile range (IQR). Other variables are presented as categorical variables using percentages. We conducted mixed-effects logistic regression analysis using the presence or absence of severe work functioning impairment as the dependent variable and subjective evaluation of the physical work environment or the number of non-conformities to the recommendations for work environments while working at home as independent variables, with the prefecture of residence as the random effect. To adjust for potential confounders, we used sex, age, education, job type, equivalent income, company size, frequency of working from home, change in working hours compared with pre-2019, one-way commute time, and cumulative infection rate by prefecture as covariates.

All statistical analyses were conducted using Stata (Stata Statistical Software: Release 14.2; StataCorp LLC, TX, USA). A *p*-value less than 0.05 was considered statistically significant.

## Results

Of a total of 33,302 responses, 6,266 cases were excluded: 215 cases were judged to have provided fraudulent responses by the company in charge of conducting the survey, and 6,051 cases met the exclusion criteria after tabulation. Of the remaining 27,036 cases, respondents who met the inclusion criteria (those worked from home at least 1 day a month) were selected, providing 5,760 cases for analysis. A flowchart showing the inclusion and exclusion of participants is shown in Figure 1.

**Figure 1.**
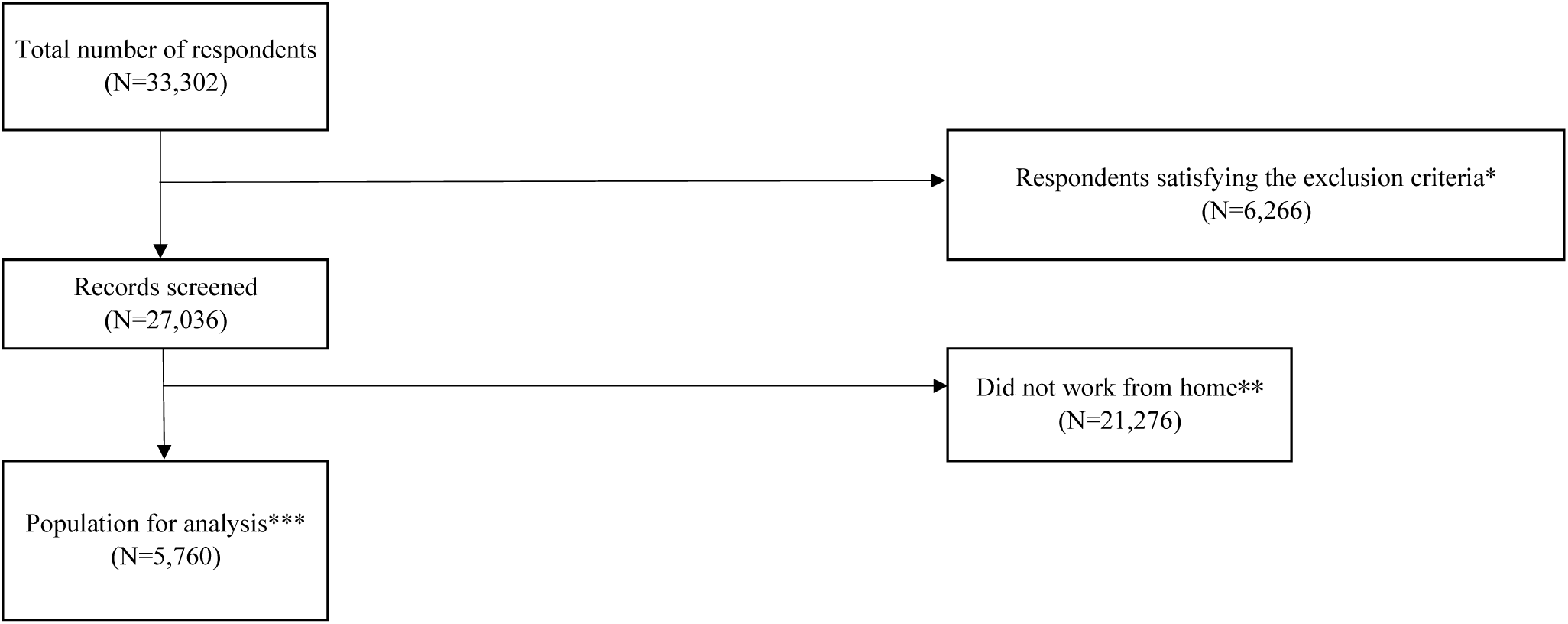
Flow chart of participants in the study *Respondents who had extremely short response times, were shorter than 140 cm tall, weighed less than 30 kg, or provided inconsistent responses to multiple identical questions. **Respondents who answered “hardly ever” to the question inquiring about the frequency of working from home *** Respondents who work from home at least 1 day a month

The demographic and sociological characteristics of the analyzed population are shown in Table 1. A total 3,361 (58%) were male and the median age was 50 years (IQR: 42–57). Of the total population, 4,052 (70%) were desk workers and 2,790 (48%) telecommuted four or more days per week. The overall median WFun score was 12 (IQR: 7–20) and severe work functioning impairment (WFun≥21) was observed in 1,309 workers (23%). There were no missing data because the survey was designed such that all responses were mandatory.

**Table 1.**
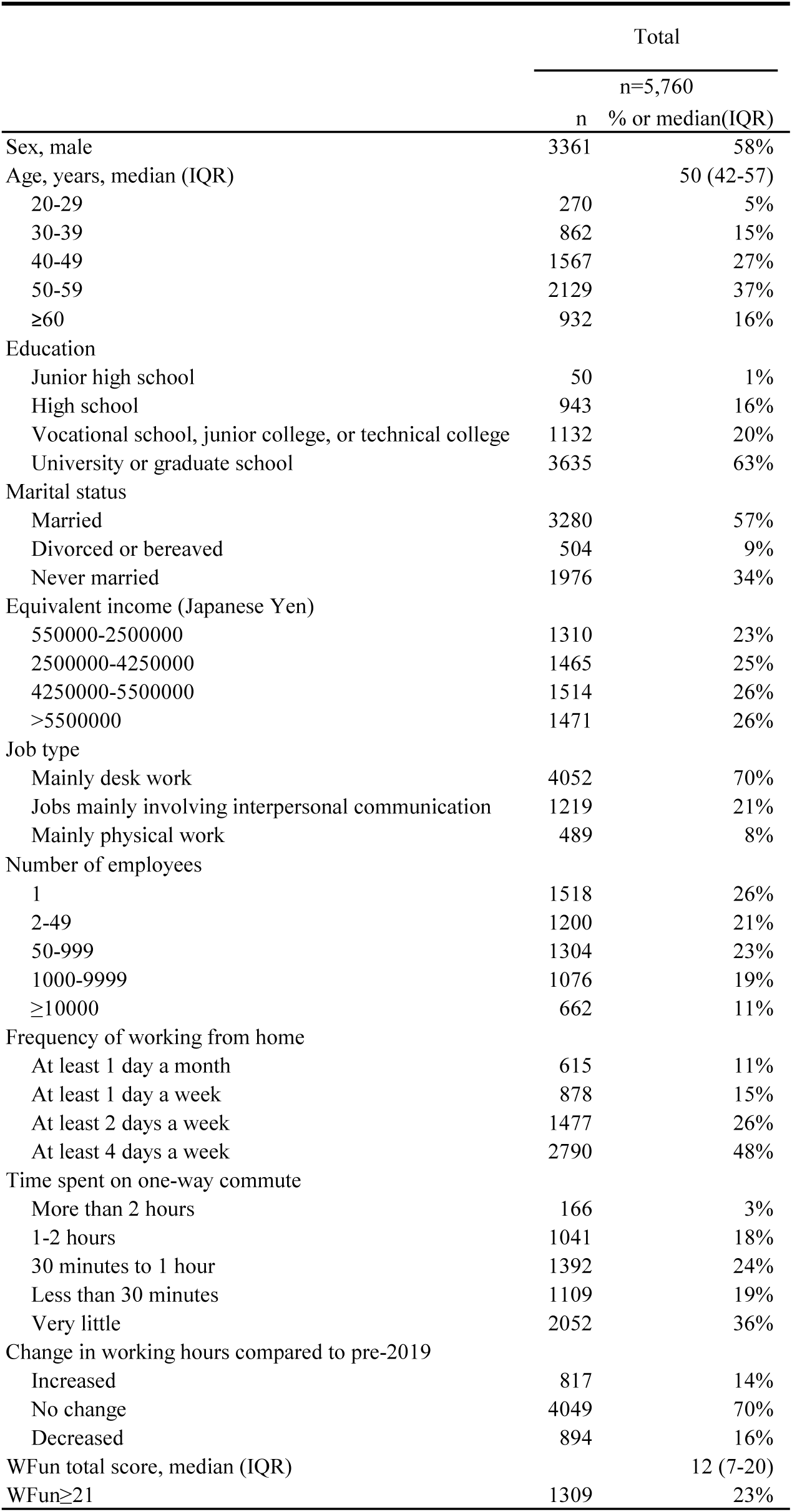
Participants’ demographic and sociological characteristics

The associations of severe work functioning impairment with workers’ physical work environment while working from home are shown in Table 2. There was no significant association of severe work functioning impairment with the item “I work at a desk/chair for office use” (adjusted odds ratio [OR]: 0.93, 95% CI: 0.82–1.06, p=0.27). For all other items, the OR of severe work functioning impairment was significantly higher among those who worked from home in an unfavorable work environment. The highest OR of work functioning impairment was for a “No” response to “There is enough light to do my work” (aOR: 2.02, 95%CI: 1.73–2.35, p<0.01). The results of Model 1, which was adjusted only for sex and age, and Model 2, which was adjusted for other potential confounders, were in a similar direction.

**Table 2.**
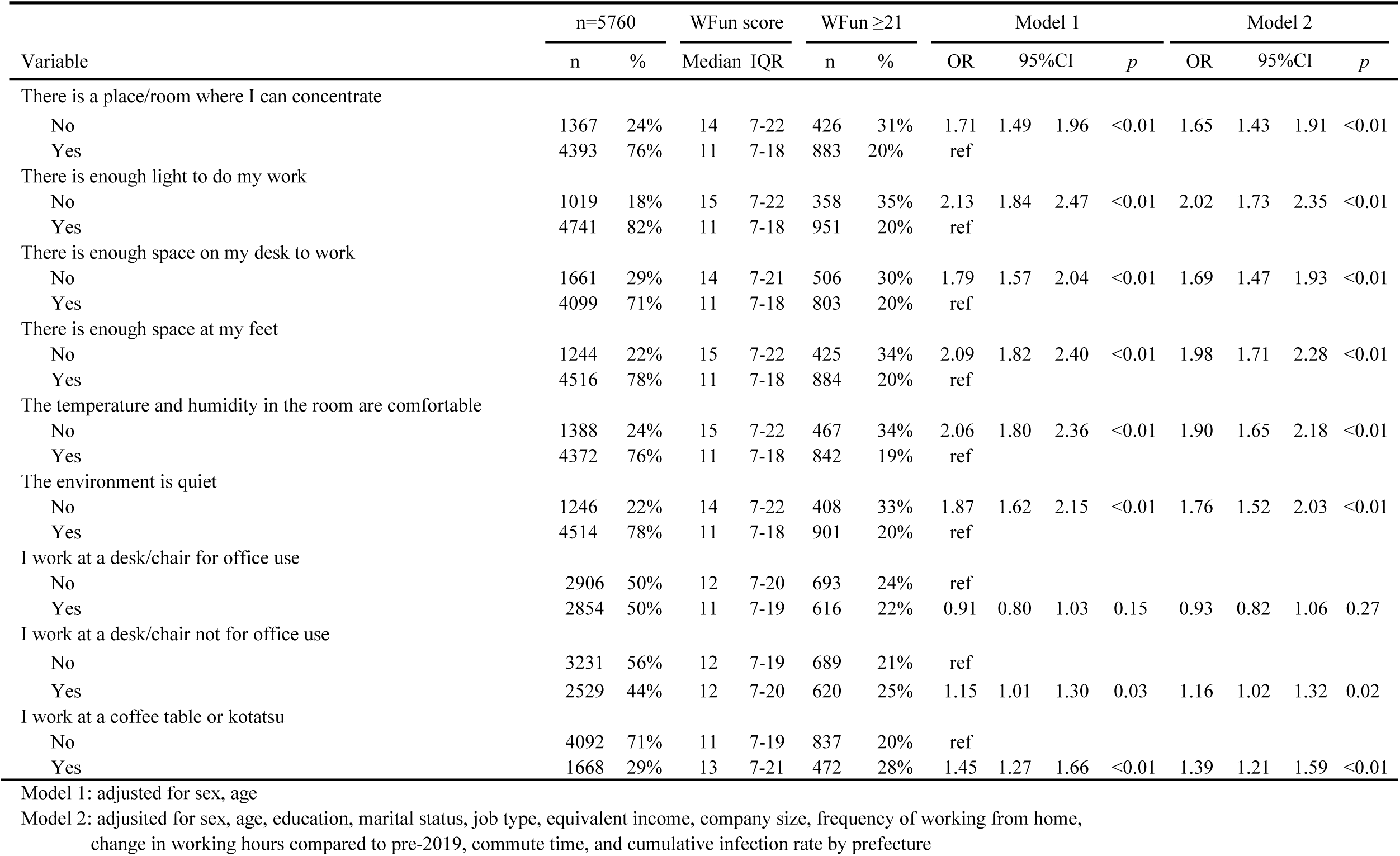
Association between workers’ physical work environment while working from home and severe work functioning impairment

Table 3 shows the relationship between severe work functioning impairment and the number of non-conformities (out of seven) to recommendations for work environments. From the nine items in Table 2, we excluded “I work at a desk/chair for office use” and combined responses to the items “I work at a desk/chair not for office use” and “I work at a table/kotatsu.” When we used those who indicated no non-conformities to recommendations for work environments as the reference group, the OR of severe work functioning impairment was significantly higher for any non-conformity to the seven items examined. Moreover, an increase in the number of non-conformities was correlated with an increase in the OR of severe work functioning impairment, with non-conformities to all seven items associated with the highest risk (aOR: 4.51, 95% CI: 2.86–7.11, p<0.01). The results of Model 1, which was adjusted only for sex and age, and Model 2, which was adjusted for other potential confounders, were in a similar direction.

**Table 3.** Association between the number of nonconformities to recommendations for work environments while working from home and severe work functioning impairment

| Variable | n=5760 |  | WFun score |  | WFun ≥21 |  | Model 1 |  |  |  | Model 2 |  |  |
| --- | --- | --- | --- | --- | --- | --- | --- | --- | --- | --- | --- | --- | --- |
|  | n | % | Median | IQR | n | % | OR | 95%CI | *p* |  | OR | 95%CI | *p* |
| Number of nonconformities to recommendations for work environments |  |  |  |  |  |  |  |  |  |  |  |  |  |
| 0 | 1177 | 20% | 8 | 7-14 | 159 | 14% | ref |  |  |  | ref |  |  |
| 1-2 | 2808 | 49% | 11 | 7-18 | 562 | 20% | 1.50 | 1.24 1.82 | <0.01 |  | 1.44 | 1.18 1.75 | <0.01 |
| 3-4 | 1084 | 19% | 14 | 7-21 | 312 | 29% | 2.38 | 1.91 2.95 | <0.01 |  | 2.18 | 1.75 2.72 | <0.01 |
| 5-6 | 598 | 10% | 16 | 7-23 | 232 | 39% | 3.79 | 2.98 4.80 | <0.01 |  | 3.46 | 2.71 4.42 | <0.01 |
| 7 | 93 | 2% | 20 | 11-27 | 44 | 47% | 5.31 | 3.40 8.28 | <0.01 |  | 4.51 | 2.86 7.11 | <0.01 |

Model 1: adjusted for sex, age
Model 2: adjusted for sex, age, education, marital status, job type, equivalent income, company size, frequency of working from home, change in working hours compared to pre-2019, commute time, and cumulative infection rate by prefecture

## Discussion

To our knowledge, this is the first study to identify an association between the physical work environment while working from home and work functioning impairment. We found that non-conformity to any of the recommendations for work environments examined in this study, except for “I work at a desk/chair for office use,” was associated with work functioning impairment. Additionally, there was a dose-response relationship between the number of non-conformities to recommendations for work environments and risk of work functioning impairment.

We found that non-conformity to any of the recommendations for work environments while working from home examined in this study, except for “I work at a desk/chair for office use,” was associated with work functioning impairment. The work environment items examined in this study are traditionally recommended for health and safety management when working at a desk (13,14,15). These included items related to the quality of the surrounds, such as temperature, humidity, and quietness of the workplace; those linked to back pain, stiff shoulders, and musculoskeletal strain; and those for concentration and performance. Comfortable temperature and humidity are expected to improve workers’ concentration and performance. In addition, items related to work posture, such as the choice of a desk and chair suitable for work and space on the desk and at the feet, are thought to reduce musculoskeletal strain during work. In addition, managing proper work posture, temperature, humidity, and illumination will help prevent visual display terminals syndrome. Setting up these environments is expected to help prevent or reduce health problems and reduce work functioning impairment while working from home.

Furthermore, our findings suggest that there is a dose-response relationship between the number of non-conformities to recommendations for work environments while working from home and work functioning impairment. In other words, no one specific item among the seven items examined in this study had a positive impact on productivity; rather, adapting as much of the work environment as possible to recommendations for work environments while working from home had an increasingly positive impact on productivity. Interestingly, only 20% of workers indicated that they conformed to all seven items examined in this study, suggesting the difficulty of creating an optimal work environment while working from home. For example, purchasing and installing new desks and chairs suitable for working from home can be a financial burden and a housing space problem. Further, adequate management of room temperature and humidity is linked to issues such as insulation of the dwelling and the purchase and electricity costs of air conditioning. Based on the results of a previous survey and our present findings, we can infer that the methods and systems for checking and improving the working environment while work from home are still lacking and that they are not being managed within companies (16). To increase productivity when working from home, both companies and workers need to account for each of these individual items when creating a work environment to ensure that as many items conform to the recommendations as possible.

There is currently no consistent evidence on the impact of working from home on the health and productivity of workers, as it varies depending on the worker’s situation (20). While working from home has its advantages, including improved work-life balance and reduced burden of commuting, it also has disadvantages, including the psychological burden of a lack of separation between living and working spaces and time, and the lack of communication with colleagues. Furthermore, as shown in this study, the impact of various aspects of the physical work environment while working from home can be significant. In fact, the number of conformities to recommendations for work environments examined in this study varied among workers. Therefore, to understand the impact of working from home on the health and productivity of workers, it is necessary to continue to study differences in the physical work environment among individuals conducting work from home.

Working from home has been suggested to be associated with sickness presenteeism, a condition in which individuals work while experiencing health problems. For example, the use of properly designed desks and chairs can affect the health of workers with musculoskeletal disorders and chronic pain, important problems that cause sickness presenteeism (21,22). Further, a study has suggested that removing barriers to work, such as commuting and going to the office, is also associated with the occurrence of sickness presenteeism (23). Improving workers’ work environment while working from home is important for reducing the negative health outcomes associated with working from home, improving presenteeism, and increasing productivity. Many companies and workers who have never had experience working from home are now suddenly being forced to do so, without appropriate preparation. As the need to telework continues, the associated risks to health and presenteeism may become more apparent, indicating the need for urgent measures to improve the work environment for those working from home.

This study has several limitations. First, selection bias was unavoidable because the study was a survey of Internet monitors. To reduce potential bias, recruitment was conducted by sampling by occupation and gender in each region according to the infection rate. To understand the characteristics of the participants in this study, we compared our findings with those from national and occupational surveys that use various batteries (17). A previous study that used WFun to examine 33,985 workers from a general company showed that 20% had severe work functioning impairment (24). Given that our study protocol found that 21% of the entire study population has severe work functioning impairment (17), we concluded that our present study population was relatively unbiased.

Second, we relied on respondents’ self-assessment of their physical environment while working from home, but did not examine the actual physical environments. Therefore, there may be discrepancies with objective evaluation. However, because we inquired about the physical environment, the possibility of erroneous answers is low.

Third, since this study was a cross-sectional study, it is impossible to determine the causal relationship between the exposure factors and outcome. However, we think it is unlikely that individuals with severe work functioning impairment would choose to create a poor working environment. For example, a person with back pain is unlikely to actively choose a small space or an ill-fitting desk environment. Therefore, there is little possibility of reverse causality.

Finally, in addition to the exposure factors examined in this study, there may be other environmental factors that can affect health and productivity while working from home. Some examples are living with a family member who needs care, inadequate telecommunication speed, and reduced physical activity.

In conclusion, individuals who work in poor work environments while working from home may exhibit work functioning impairment during a period of rapid change owing to the COVID-19 pandemic. As work environments become more diverse, it is important for both companies and individual workers to create a work environment that prevents negative health outcomes and improves productivity while working from home.

## Data Availability

Due to the nature of this research, the participants of this study did not agree to their data being shared publicly, and hence, supporting data is not available.

